# Health-related quality of life of female sex workers living with HIV in South Africa: a cross-sectional study

**DOI:** 10.1101/2021.06.07.21258225

**Authors:** Linwei Wang, David W. Dowdy, Carly A. Comins, Katherine Young, Mfezi Mcingana, Ntambue Mulumba, Hlengiwe Mhlophe, Claire Chen, Harry Hausler, Sheree R. Schwartz, Stefan Baral, Sharmistha Mishra, the Siyaphambili Study team

**Author notes:** **Corresponding Author:** Sharmistha Mishra, MD, MSc, PhD, MAP-Centre for Urban Health Solutions, St. Michael’s Hospital, Unity Health Toronto, University of Toronto, 209 Victoria St, Toronto, ON M5B 1T8, E. **E-mail addresses of authors:** LW DWD CAC KY MM NM HM CC HH SRS SB SM.

## Abstract

**Introduction:** Health-related quality of life (HRQoL) is an important HIV outcome beyond viral suppression. However, there are limited data characterizing HRQoL of key populations including female sex workers (FSW) living with HIV.

**Methods:** We used baseline data (June 22, 2018-March 23, 2020) of FSW who were diagnosed with HIV and enrolled into a randomized trial in Durban, South Africa. HRQoL information was collected by a generic preference-accompanied tool with five domains (EQ-5D), and summarized into a single score (range 0-1) which represents health utility. We employed multivariable beta regression models to identify determinants of HRQoL and to estimate subgroup-specific HRQoL score. Using external estimates of life expectancy and population size, we estimated the number of quality adjusted life years reduced among FSW living with HIV in South Africa associated with violence and drug use.

**Results:** Of 1363 individuals (mean age: 32.4 years; mean HRQoL score: 0.857) in our analysis, 62.6% used drugs, 61.3% experienced physical or sexual violence, and 64.6% self-reported taking antiretroviral treatment (ART). The following were associated with a reduction in the average marginal HRQoL score: older age (per decade: 0.018 [95% confidence interval (CI): 0.008, 0.027]), drug use (0.022 [0.007, 0.036]), experience of violence (0.024 [0.010, 0.038]), and moderate (vs. no) level of internalized stigma (0.023 [0.004, 0.041]). Current ART use was associated with a 0.015-point (−0.001, 0.031) increase in the HRQoL score. The estimated mean (95%CI) HRQoL scores ranged from 0.838 (0.816, 0.860) for FSW who used drugs, experienced violence, and were not on ART; to 0.899 (0.883, 0.916) for FSW who did not use drugs nor experience violence and were on ART. Our results can be translated into a reduction in 37,184 and 39,722 quality adjusted life years related to drug use and experience of violence, respectively in South Africa.

**Conclusions:** These results demonstrate the association of ART with higher HRQoL among FSW and the need to further address structural risks including drug use, violence, and stigma. Population-specific estimates of HRQoL score can be further used to calculate quality-adjusted life years in economic evaluations of individual and structural interventions addressing the needs of FSW living with HIV.

**Clinical Trial Registration:** NCT03500172 (April 17, 2018)

## INTRODUCTION

Globally, there has been remarkable progress towards the UNAIDS 90-90-90 HIV testing and treatment targets. By the end of 2019, 81% of people living with HIV (PLHIV) knew their status, of whom 83% were on antiretroviral treatment (ART), and 88% of those on ART were virally suppressed(1). Although much work remains to improve the cascade of HIV care in order to achieve the new 95-95-95 targets by 2030(2), there is growing recognition of the importance of patient-centered chronic care to improve patient-reported outcomes such as health-related quality of life (HRQoL)(3, 4). Some have proposed that good HRQoL should be explicitly included as the ‘fourth’ target as part of the HIV care cascade(4).

HRQoL is a multidimensional concept defined as a person’s subjective perception of the impact of ill health on daily life and includes physical, psychological and social functioning(5). Most existing studies on HRQoL among PLHIV have identified CD4 count, gender, age, and ART use as common determinants for HRQoL(6-9). The majority of these studies have been conducted in wider populations of PLHIV in high income settings(8), though some have been performed in low and middle income settings including sub-Saharan Africa(7, 8). There is a dearth of research regarding HRQoL in key populations living with HIV in high prevalence settings, including among female sex workers (FSW).

In South Africa, FSW have a high burden of HIV(10). Approximately 90,000 FSW in South Africa are living with HIV, representing nearly 60% of all women engaged in sex work(10). FSW living with HIV face a wide range of challenges, including HIV progression, comorbidities, drug use, stigma, and other social and structural vulnerabilities(3, 8, 9) resulting in suboptimal treatment uptake and viral suppression(11-13). Data, although limited, suggest that less than 40% of FSW living with HIV are virally suppressed(11). However, it is unclear how these challenges have shaped FSW’s self-perception of their well-being. A meta-analysis across 31 studies in low and middle income settings found highly prevalent mental health conditions among FSW, including depression, anxiety, post-traumatic stress disorder, and suicidal thoughts and attempts(14). However, these studies examined FSW in general, instead of PLHIV, and with a focus on mental health rather than HRQoL(14).

HRQoL as a multidimensional concept can also be summarized into a single metric of health utility, which is important for economic evaluations of HIV interventions(6, 7, 15). For example, preference-accompanied measures of HRQoL such as the EQ-5D (captures self-rated information in five domains - mobility, self-care, ability to do usual activities, pain/discomfort, and anxiety/depression, accompanied by a set of health state utility values) are often summarized into a single score representing health utility, and used to calculate quality-adjusted life years (QALYs) to be applied in cost-utility analyses(15-19). A review of the economic evaluation literature on HIV interventions in sub-Saharan Africa reveals that the weights assigned to QALYs often have an insufficient evidence due to lack of data on setting- and population-specific measures(7, 20).

To address these gaps, we aimed to examine HRQoL across the five EQ-5D domains, estimate HRQoL scores which represent health utilities, and identify characteristics associated with HRQoL scores among FSW living with HIV in Durban, South Africa.

## METHODS

### Study design and subjects

The current analysis uses baseline data from an adaptive randomized intervention trial (Siyaphambili study) involving FSW in Durban, South Africa(11). Cisgender women who were 18 years and older, selling sex as their main source of income, residing in Durban, and diagnosed with HIV at least 6 months prior to study enrollment were eligible. Full details of the study have been described elsewhere(11) and in **Appendix 0**.

Following informed consent and enrollment, participants provided whole blood samples which were used to assess baseline CD4 count and viral load by the South African National Health Laboratory Services per national guidelines. Individuals completed an interviewer-administered questionnaire that elicited information including socio-demographics, personal/sexual history, HIV care, drug use, and HRQoL.

All FSW recruited between June 22, 2018 and March 23, 2020 who completed their baseline questionnaire were included. Individuals were excluded if their responses to the primary outcome (described below) were missing.

### Measures

HRQoL information was collected using the generic preference-accompanied measure - EQ-5D-3L(16). EQ-5D-3L measures HRQoL in five domains (mobility, self-care, ability to do usual activities, pain/discomfort, and anxiety/depression), and each domain is measured with three levels (1=no problems, 2=some/moderate problems, and 3=extreme problems). The answers can be assembled into a 5-digit health state reflecting the score on each dimension, resulting in a total of 243 plausible health states (3^5). EQ-5D also contains a separate question which asks participants to directly rate their health state from the best (score of 100) to the worst (score of 0) (referred to as EQ visual analogue scale (EQ-VAS)). EQ-5D has been used to measure HRQoL of the wider population and PLHIV across settings including South Africa(7, 8, 21, 22). EQ-5D is also a guideline-recommended instrument to derive health utility measures(15, 23).

Our primary outcome was a summary HRQoL score, representing health utility. The HRQoL score was calculated using the 5-digit health states combined with valuations of the health states from the wider population (value set). A wider population value set is used to reflect the preferences of local taxpayers and potential receivers of healthcare(16). We used the value set obtained from the population in Zimbabwe(21). The calculated HRQoL score ranges between 0-1, where 0 represents a state equivalent to death and 1 represents perfect health.

Our secondary outcomes included EQ-VAS, each of the EQ-5D-3L domains, and a binary variable indicating no reported problem on all five EQ-5D-3L domains (perfect health).

We defined an a priori set of covariates based on evidence from the literature regarding their associations with HRQoL(7, 9, 24). These covariates included sociodemographics, clinical features and treatment of HIV, drug use and other social and structural factors. Specifically, we considered age, race, education, ART experience, CD4 count, viral load, drug use in the past 30 days, lifetime injection drug use, experience of homelessness in the past 6 months, lifetime experience of physical or sexual violence, experience with stigma related to HIV or sex work, and internalized stigma related to sex work. Details of each variable are shown in **Table 1** and **Appendix A1**.

**Table 1.**
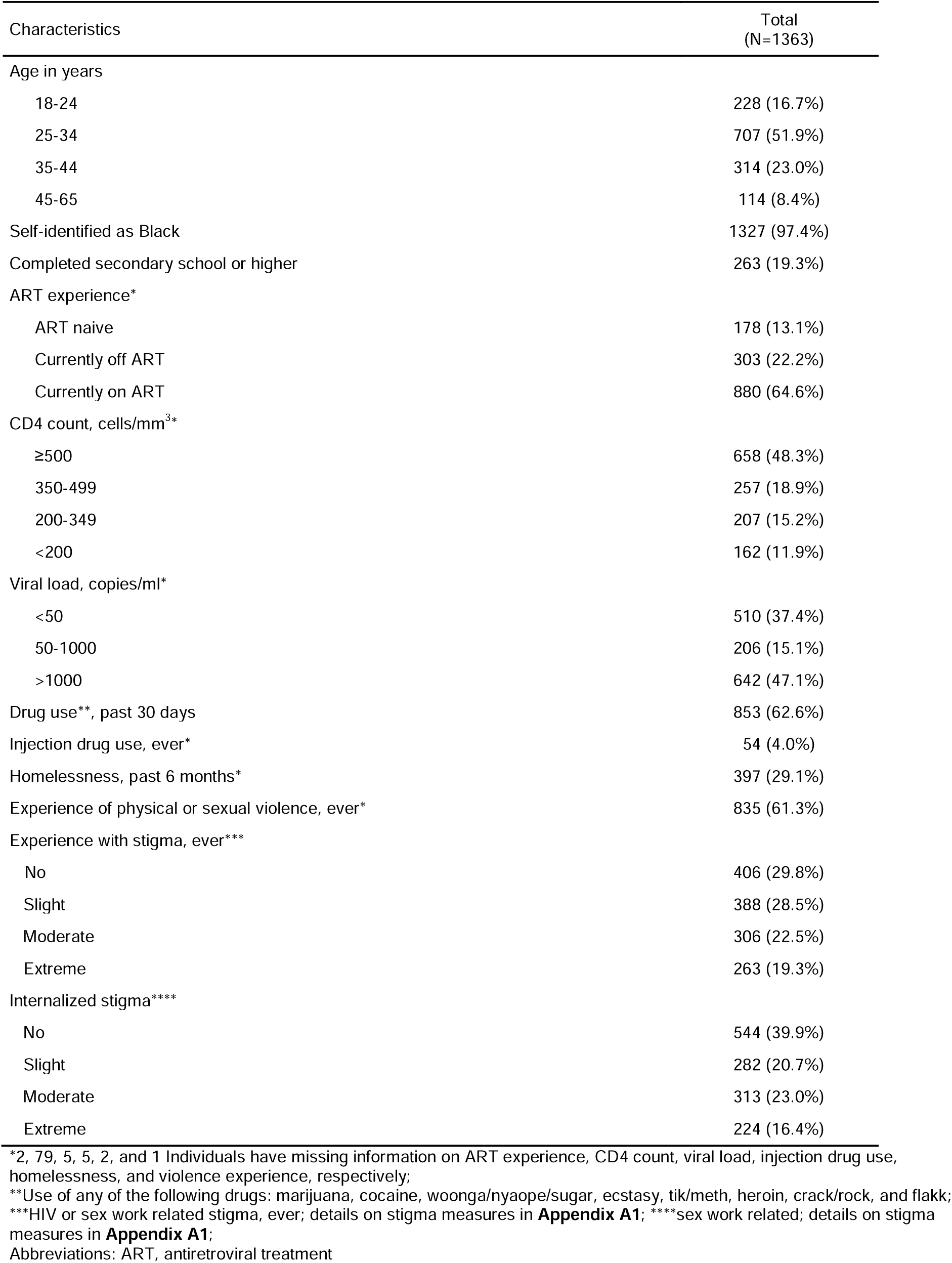
Characteristics of female sex workers living with HIV in Durban, South Africa, 2018-2020.

| Characteristics | Total<br>(N=1363) |
| --- | --- |
| Age in years |  |
| 18-24 | 228 (16.7%) |
| 25-34 | 707 (51.9%) |
| 35-44 | 314 (23.0%) |
| 45-65 | 114 (8.4%) |
| Self-identified as Black | 1327 (97.4%) |
| Completed secondary school or higher | 263 (19.3%) |
| ART experience* |  |
| ART naive | 178 (13.1%) |
| Currently off ART | 303 (22.2%) |
| Currently on ART | 880 (64.6%) |
| CD4 count, cells/mm<sup>3</sup>* |  |
| ≥500 | 658 (48.3%) |
| 350-499 | 257 (18.9%) |
| 200-349 | 207 (15.2%) |
| <200 | 162 (11.9%) |
| Viral load, copies/ml* |  |
| <50 | 510 (37.4%) |
| 50-1000 | 206 (15.1%) |
| >1000 | 642 (47.1%) |
| Drug use**, past 30 days | 853 (62.6%) |
| Injection drug use, ever* | 54 (4.0%) |
| Homelessness, past 6 months* | 397 (29.1%) |
| Experience of physical or sexual violence, ever* | 835 (61.3%) |
| Experience with stigma, ever*** |  |
| No | 406 (29.8%) |
| Slight | 388 (28.5%) |
| Moderate | 306 (22.5%) |
| Extreme | 263 (19.3%) |
| Internalized stigma**** |  |
| No | 544 (39.9%) |
| Slight | 282 (20.7%) |
| Moderate | 313 (23.0%) |
| Extreme | 224 (16.4%) |

\*2, 79, 5, 5, 2, and 1 Individuals have missing information on ART experience, CD4 count, viral load, injection drug use, homelessness, and violence experience, respectively;
\*\*Use of any of the following drugs: marijuana, cocaine, woonga/nyaope/sugar, ecstasy, tik/meth, heroin, crack/rock, and flakk; \*\*\*HIV or sex work related stigma, ever; details on stigma measures in **Appendix A1**; \*\*\*\*sex work related; details on stigma
measures in **Appendix A1**;
Abbreviations: ART, antiretroviral treatment

### Statistical analysis

First, we examined the descriptive characteristics of the study sample at baseline; assessed the distribution of our primary and secondary measures of HRQoL; and compared the differences in the HRQoL score, and of EQ-VAS, by covariates. Wilcoxon rank-sum and Kruskal-Wallis tests were used to compare the median values in outcomes across binary variables and other categorical variables, respectively.

Second, we employed multivariable beta regression models with a complementary log-log link and fixed dispersion to investigate associations between covariates of interest and the HRQoL score(25). Beta regression models provide a flexible approach to modeling health utility values which are truncated and skewed(25). We provide details on model selection in **Appendix A2**. We present the results as average marginal effects along with 95% confidence intervals (CIs), which reflect the magnitude of association between a one-unit change in covariates and the mean HRQoL score(26).

Using the fitted model, we estimated the mean and 95% CIs of HRQoL score for subgroups of FSW defined by identified determinants of HRQoL score, setting the rest covariates at the sample average values. In addition, assuming an average remaining life expectancy of 30 years(27), and an estimated 90,000 FSW living with HIV in South Africa(10), using the observed prevalence of drug use and experience of violence, we estimated the number of QALYs related to drug use and experience of violence, respectively in South Africa.

Third, we performed a set of multivariable regression analyses to examine the relationship between the covariates of interest and our secondary outcomes. Multivariable linear regression was used to model EQ-VAS. Multivariable logistic regression was used to model each of the EQ-5D-3L domains, and the perfect health state, separately. We grouped each EQ-5D-3L domain into two levels (reporting moderate/extreme vs. no problems).

Lastly, to examine the sensitivity of our results to the value set used, we computed a new HRQoL score using the value set obtain from the wider population in UK(22). We repeated the multivariable beta regression analysis to examine the relationship between covariates of interest and the new HRQoL score.

All statistical analyses were executed in R version 3.6.2. R package ‘eq5d’ was used to calculate the HRQoL score(28), ‘betareg’ was used for beta regression analysis(25), and ‘margins’ was used to estimate average marginal effects(26).

## RESULTS

Of 1363 individuals included in our analysis, the majority (97.4%) self-identified as black, and the mean age was 32.4 years (standard deviation: 8.0; median: 31.0; interquartile range: 26.6-36.7). We excluded 28 (2.0%) individuals because they were missing the primary outcome (**Appendix Figure A3.1** shows the details on exclusion). Overall, 853 individuals (62.6%) reported using drugs in the past 30 days, 397 (29.1%) experienced homelessness in the past six months, 835 (61.3%) ever experienced physical or sexual violence, and 1199 (88.0%) reported either experience with stigma (n=957, 70.2%) or internalized stigma (n=819, 60.1%). All individuals were diagnosed with HIV prior to study entry, of whom 178 (13.1%) self-reported being ART naïve while 880 (64.6%) reported being on ART, and 303 (22.2%) reported prior, but non-current ART use at time of enrollment. According to blood tests conducted at study entry, 510 (37.4%) individuals were virally suppressed (<50 copies/ml). **Table 1** provides details on sample characteristics.

Regarding the five domains of EQ-5D, the majority of individuals reported no problems in mobility (95.7%), self-care (99.0%), and usual activity (96.9%); while 32.9% and 40.7% reported at least some problems in pain/discomfort, and in anxiety/depression, respectively (**Figure 1A**). Of the 243 plausible health states summarized across EQ-5D domains, we observed 47; the 12 most frequent health states represented more than 95% of all observations (**Figure 1B**). Individuals with a perfect health state accounted for more than half (52.5%) of the sample (**Figure 1B**). The HRQoL score was heavily left-skewed, ranging from 0.233 to 1, with a median of 1 [interquartile range: 0.787,1] and mean of 0.857 (standard deviation: 0.187) (**Figure 1C**). EQ-VAS ranged from 0 to 100, with a median of 52.0 [50.0, 70.0] and mean of 57.7 (21.9) (**Figure 1D**).

**Figure 1.**
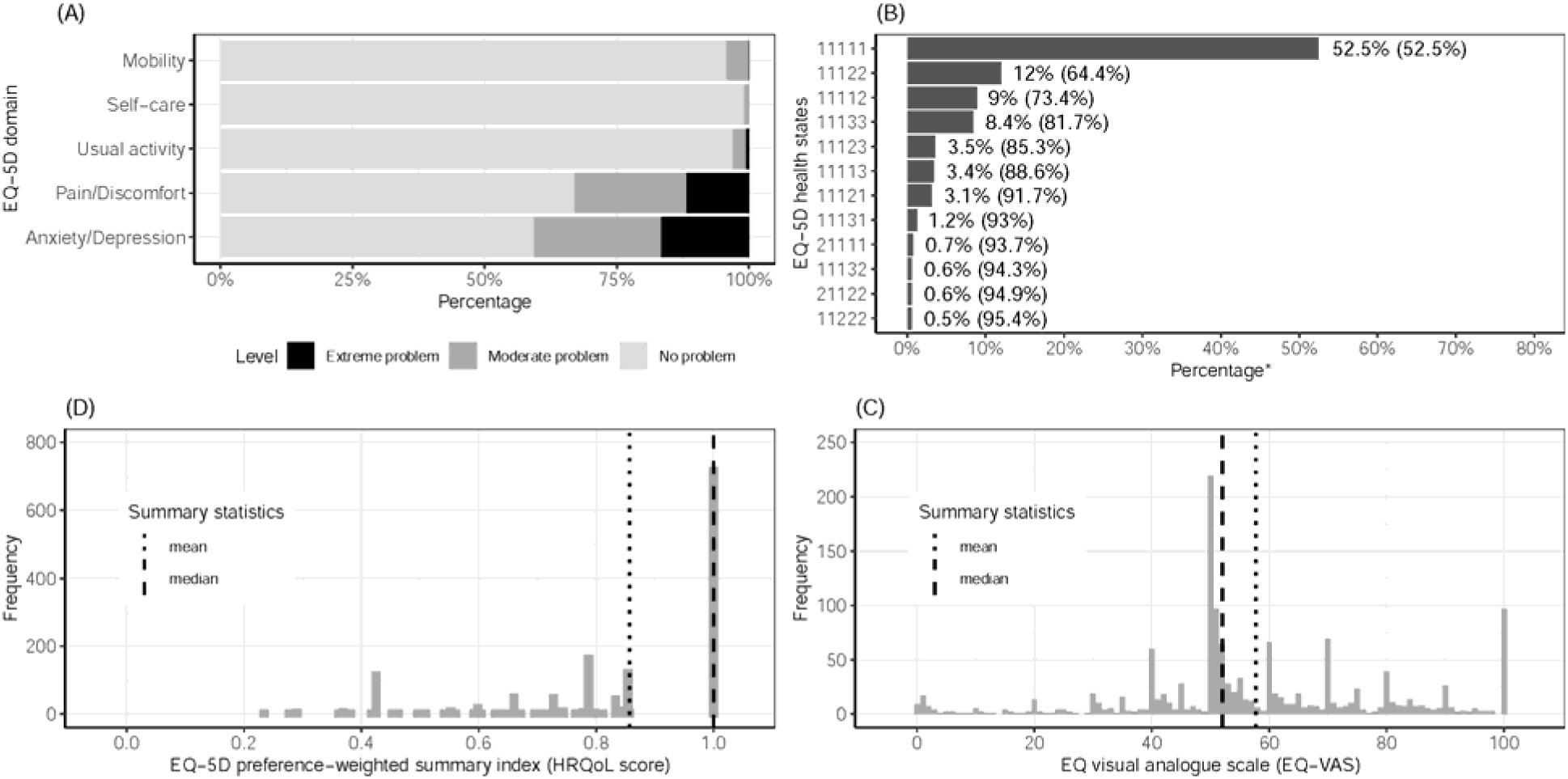
Health-related quality of life profile of female sex workers living with HIV in Durban, South Africa, 2018-2020. (A) Proportion of women reporting problems on each EQ-5D-3L domain. (B) Most frequent EQ-5D-3L health states. For example, health state 11123 refers to no problems in mobility, self-care and usual activity, moderate problems in pain/discomfort, and extreme problems in anxiety/depression. *Numbers in the bracket denote the cumulative percentage of health states from top to bottom. (C) Distribution of EQ-5D-3L preference-weighted summary index (HRQoL score, representing health utility), in which 1 denotes no problems in any of the five domains and 0 denotes a health state equivalent to death. (D) Distribution of EQ-5D visual analogue scale, in which participants are asked to rate their quality of life on a “thermometer” from 0 (equivalent to death) to 100 (perfect health). Abbreviations: EQ-5D-3L, EuroQoL five dimensions, three levels; HRQoL, health-related quality of life.

**Table 2** presents the summary statistics of HRQoL score and EQ-VAS by sample characteristic. The median HRQoL score was higher in younger individuals (1.00 vs. 0.854, comparing ages 18-34 vs. 35-64, p=0.006); those identified as black (1.00 vs. 0.854, p=0.008) and reported current ART use (1.00 vs. 0.854, p=0.001). In contrast, the median HRQoL score was lower in individuals who used drugs (0.854 vs. 1.00, p<0.001), and who experienced homelessness (0.854 vs. 1.00, p=0.014) or violence (0.854 vs. 1.00, p<0.001). We did not observe a difference in HRQoL score by CD4 count nor by viral load. However, individuals with higher CD4 counts and lower viral load had higher EQ-VAS (p<0.001, **Table 2**); and those with experience of or internalized stigma had lower EQ-VAS (p<0.001, **Table 2**).

**Table 2.**
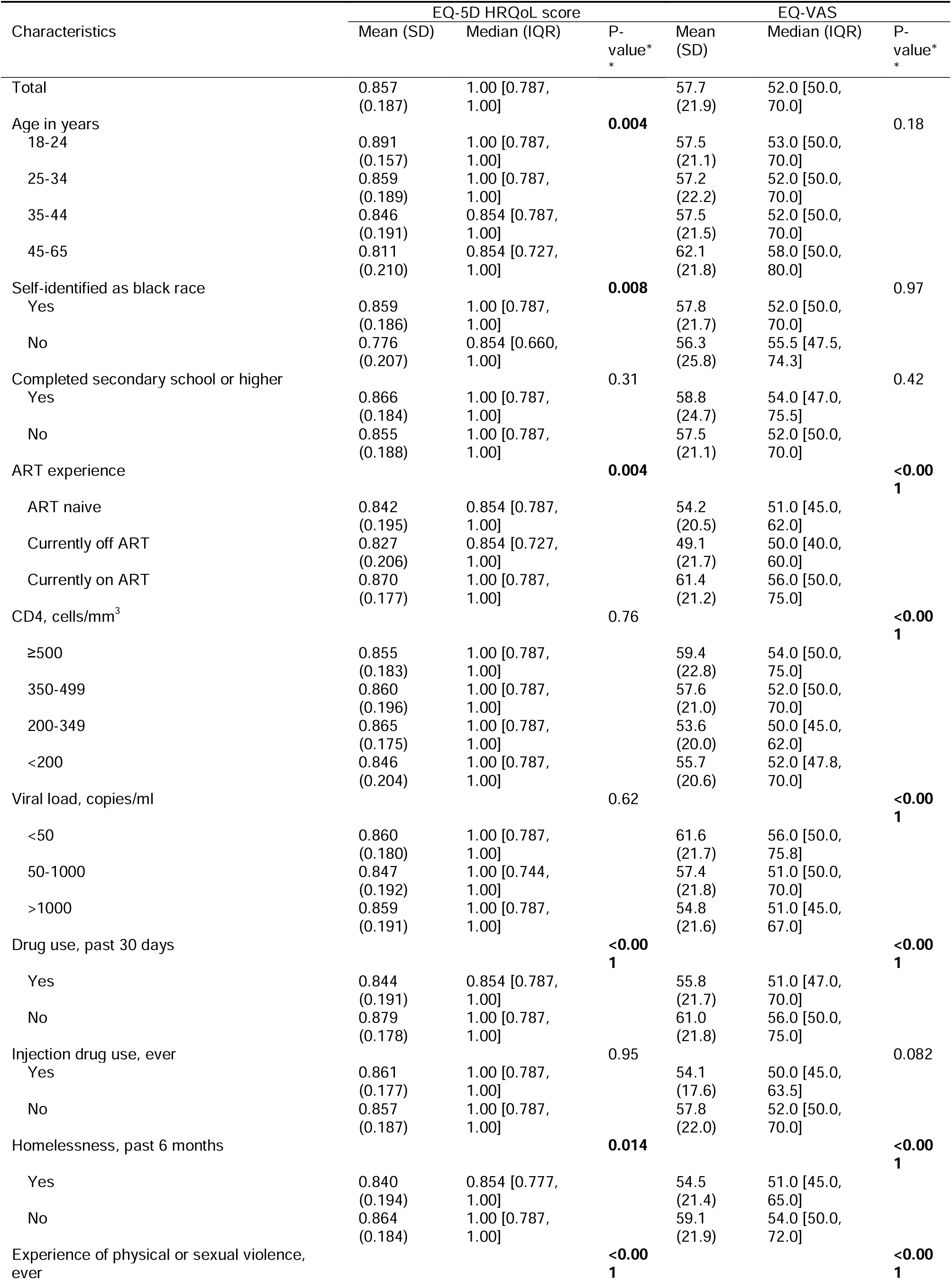
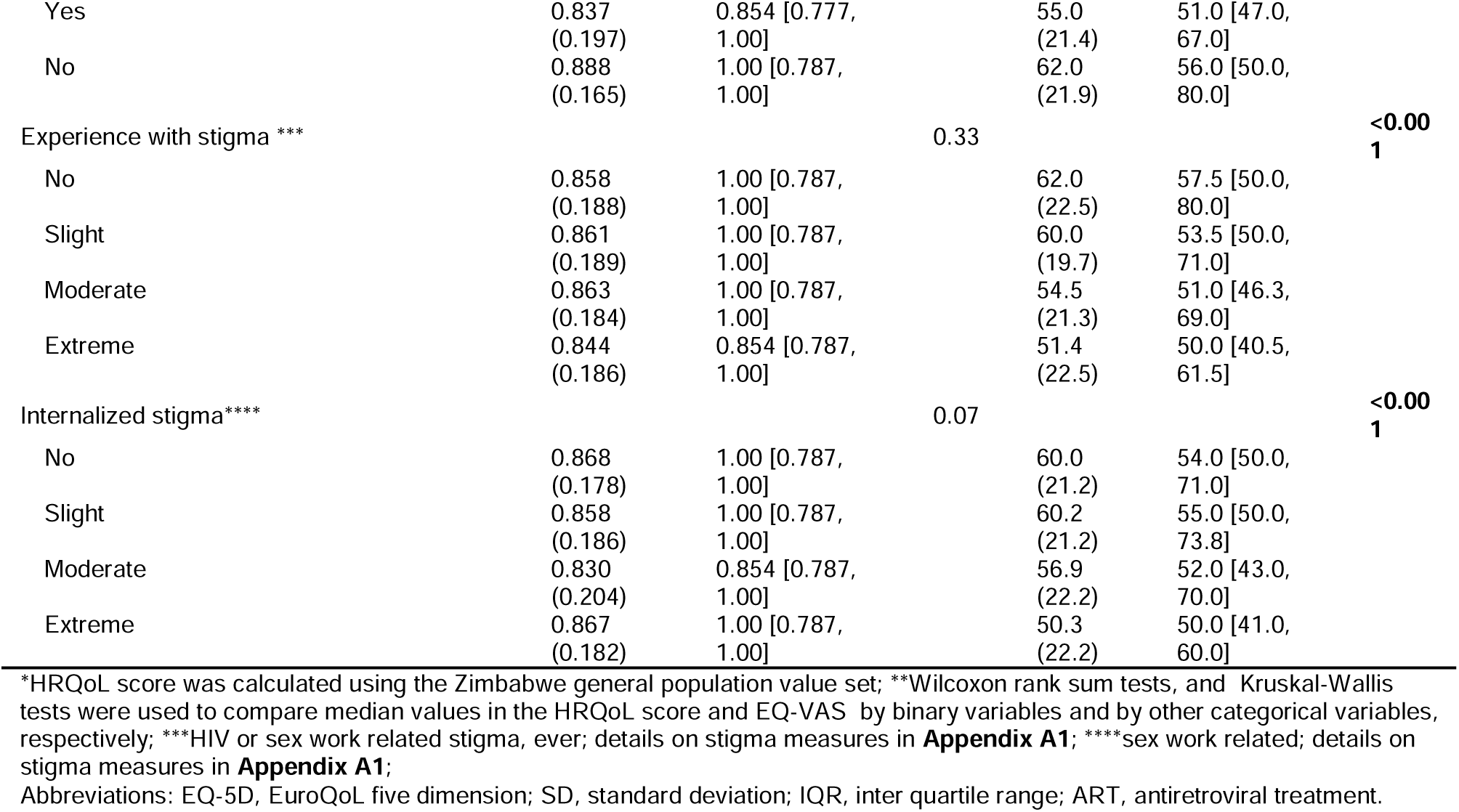
EQ-5D health-related quality of life (HRQoL) score* and visual analogue scale (EQ-VAS) among female sex workers living with HIV in Durban, South Africa, 2018-2020.

In the multivariable regression analyses, we excluded another 87 (6.4%) individuals because they had missing covariate information (**Appendix Figure A3.1**). Individuals excluded were younger, less likely to be on ART, and more likely to experience homelessness compared to those included (**Appendix Table A3.1**). We found that age (per ten-year increase), drug use, and violence experience were associated with 0.018 (95% CI: 0.008, 0.027)), 0.022 (0.007, 0.036), and 0.024 (0.010, 0.038) lower average marginal HRQoL score, respectively (**Table 3**). Current ART use was associated with 0.015 (−0.001, 0.031) higher average marginal HRQoL score (**Table 3**). We did not find a difference in HRQoL score between individuals who were ART naïve and who had previously been but were not currently on ART (results not shown; we thus combined these two groups in models). Individuals with moderate internalized stigma had 0.023 (0.004, 0.041) lower average marginal HRQoL score compared to those without; however, we did not observe a difference between those with extreme versus no internalized stigma (**Table 3**). We did not find independent associations between education, CD4 count, viral load, homelessness, or stigma experience with HRQoL score (**Table 3**).

**Table 3.**
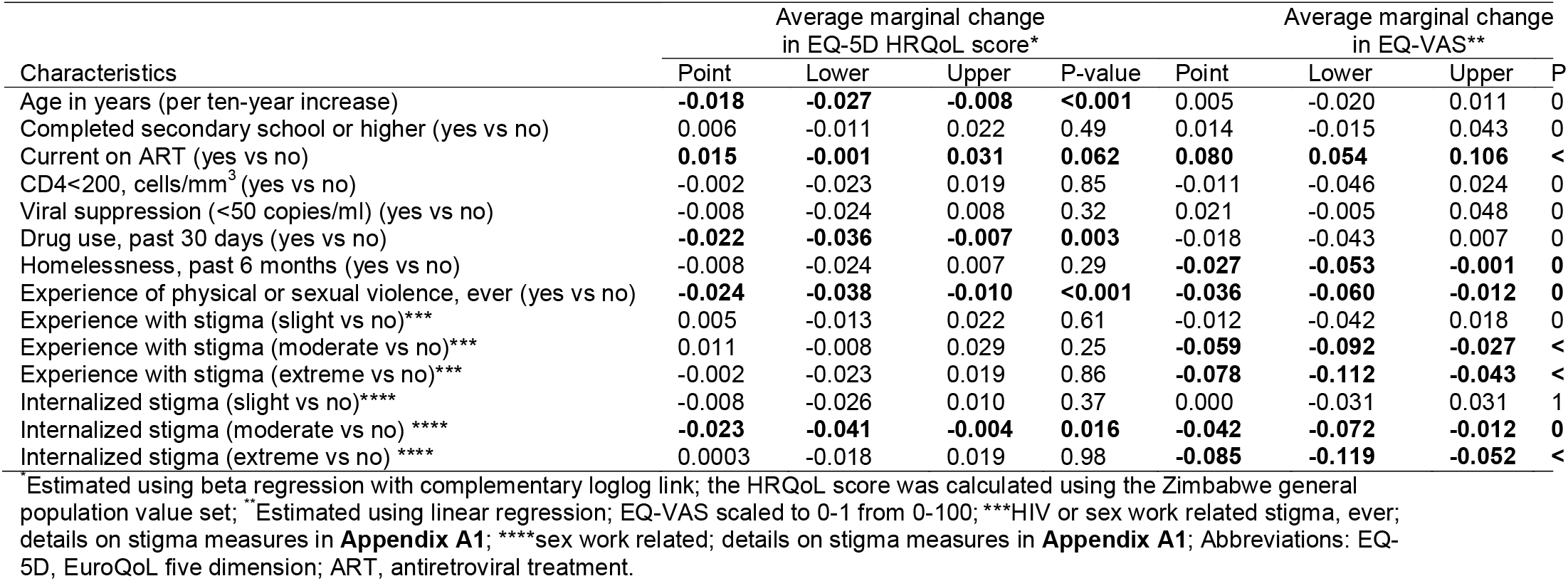
Multivariable analyses of factors associated with the EQ-5D health-related quality of life (HRQoL) score and the visual analogue scale (EQ-VAS) of female sex workers living with HIV in Durban, South Africa, 2018-2020.

Based on the fitted model, the estimated mean (95% CI) HRQoL scores were 0.870 (0.854, 0.886) for FSW living with HIV overall, 0.877 (0.862, 0.893) and 0.862 (0.843, 0.882) for those with and without current ART use, respectively. **Figure 2** shows estimates of HRQoL score for other subgroups of FSW. For example, the model estimated mean HRQoL scores of 0.838 (0.816, 0.860) for FSW who used drugs, experienced violence, and were not on ART, versus 0.899 (0.883, 0.916) for FSW who did not use drugs nor experience violence and were on ART. Our results can be translated into a reduction in 37,184 and 39,722 QALYs related to drug use and violence experience, respectively in South Africa.

**Figure 2.**
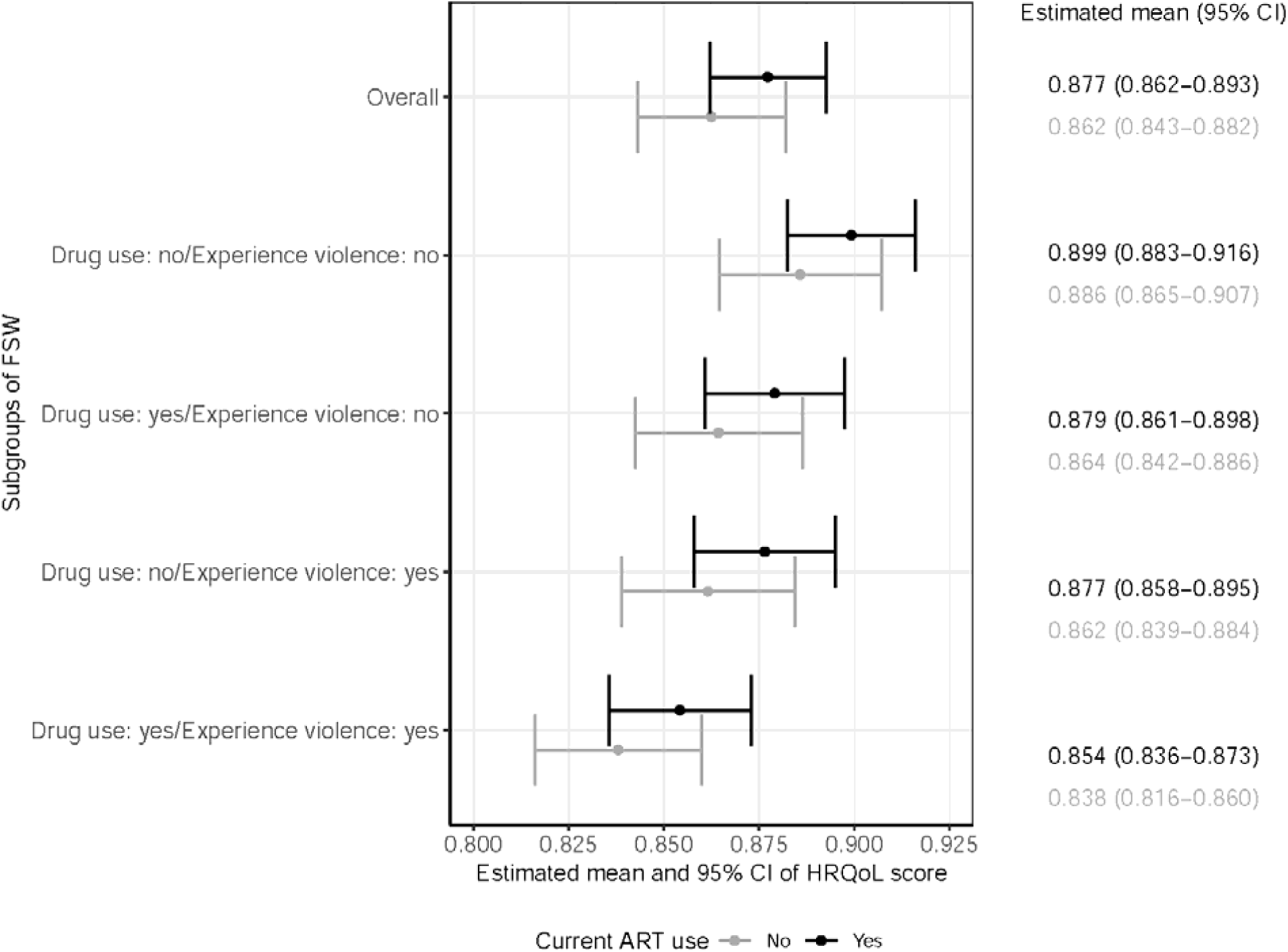
Estimated mean and 95% confidence intervals of health-related quality of life (HRQoL) score (representing health utility) for subgroups of female sex workers (FSW) living with HIV in Durban, South Africa by current antiretroviral treatment (ART) use. Values estimated using the fitted multivariable beta regression model, setting the rest of covariates (age, education, CD4 count, viral suppression, homelessness, experience with stigma and internalized stigma) at the sample average values.

Multivariable analyses of secondary HRQoL outcomes identified some similarities and differences in terms of variables associated HRQoL (**Table 3**; **Appendix Figure A3.2**). For example, age and drug use were not independently associated with EQ-VAS (**Table 3**). Homelessness was associated with lower EQ-VAS (−0.027 (−0.053, -0.001)) (**Table 3**). We observed a dose-response relationship between both stigma variables and EQ-VAS, where increasing stigma was associated with lower EQ-VAS (**Table 3**). When we analyzed the five domains of EQ-5D separately, we found homelessness was associated with increased odds of reporting any problem in mobility (1.72 (0.95, 3.09)), and in pain/discomfort (1.46 (1.11, 1.90)) ((**Appendix Figure A3.2**).

Our sensitivity analysis using a UK-based value set revealed similar results (**Appendix Table A3.2**).

## DISCUSSION

There has been limited study of HRQoL among FSW living with HIV across Sub-Saharan Africa including the estimation of health utility. Among 1363 FSW who were diagnosed with HIV and enrolled into an adaptive randomized trial via HIV prevention and treatment program for sex workers in Durban, we found older age, lack of ART use, drug use, experience of violence, and moderate (vs. no) level of internalized stigma were associated with lower HRQoL score at the time of enrollment. In addition, we present the overall, and subgroup-specific estimates of HRQoL scores, which can be used to calculate QALYs in cost-utility analysis of interventions focused on FSW living with HIV in South Africa.

The observed average HRQoL score for FSW living with HIV was 0.857 (model estimated mean of 0.870 (95% CI: 0.854, 0.886)), which is somewhat lower than an estimate from the wider population living with HIV in South Africa between 2013-2015 (0.89(24)), when evaluated using the same value set(21). In fact, the lower HRQoL score for FSW living with HIV stemmed largely from concerns in pain/discomfort (32.9% of FSW in our study reported pain/discomfort vs. 5% of wider population living with HIV in South Africa(24)) and anxiety/depression (40.7% vs. 5%(24)). Our findings are consistent with previous studies which found higher burden of mental health conditions among FSW in low and middle income settings compared to the wider population, though none of these studies directly estimated HRQoL as a measure of health utility(14). The associations between lower HRQoL and drug use, as well as experience of violence, were also consistent with this recent meta-analysis(14). Lower HRQoL associated with experience of violence may reflect pain and psychologic effects (e.g., depression, or post-trauma syndrome) of violence (29). Lower HRQoL associated with drug use may reflect self-medication as a coping strategy in the context of chronic pain or depression(30, 31), and/or reflect mental health effects of chronic use of drugs(31). The high prevalence of drug use (62.6%), and experience of violence (61.3%) among FSW living with HIV, and their associations with lower HRQoL further demonstrate the synergistic epidemics of substance abuse, violence and HIV/AIDS (SAVA syndemic) among women(29, 32); and highlight the need for integrated services to prevent and mitigate the effects of violence and drug use to help improve the well-being among sex workers.

We also observed a high prevalence of homelessness (29%) and experience of stigma or internalized stigma (88%) among FSW living with HIV in the study. Although we did not observe an independent association between experience of homelessness or stigma with HRQoL score, homelessness was associated with increased odds of reporting problems in mobility, pain/discomfort, as well as lower EQ-VAS. We also found a strong negative dose-response relationship between levels of stigma and the EQ-VAS. These findings suggest a strong negative subjective experience of stigma, even if this experience is not reflected in self-reported problems on the EQ-5D domains. Although generic preference-accompanied measures of HRQoL such as EQ-5D allow for calculation of HRQoL scores which enables comparison across groups with different diseases, these measures do not capture other aspects of HRQoL such as social relationships, spirituality, and environment (e.g., physical safety and security)(33), and may be insensitive to specific issues faced by specific populations(34).

Moreover, HRQoL score is calculated by valuing individual’s self-rated health state from a wider population’s perspective of health preference, which contrasts with EQ-VAS, reflecting purely individual’s subjective perception of health(16). Other reasons underlying the non-monotonic relationship (although not statistically significant) between stigma and self-reported problems on the EQ-5D domains may include the following: stigma may be associated with individual’s definition/perception of ‘having a problem’/tolerance levels of the problem(35); as well as individual’s tendency to provide socially desirable answers(36). Future studies using disease-specific HRQoL measures such as the WHOQOL-HIV BREF (33) and exploring various underlying types of stigma(37) may provide additional insights into the relationship between stigma and HRQoL among FSW living with HIV. Our findings of high prevalence of stigma and its association with poorer self-perceived overall HRQoL further highlight the need to evaluate interventions to combat intersectional stigma (e.g., the convergence of multiple stigmatized attributes) for key populations(38).

Despite heterogeneity in HRQoL across subgroups of FSW living with HIV, we found that current ART use was independently associated with higher HRQoL. These results are consistent with previous studies among the wider population(7, 15, 24), including studies with longitudinal design which found improvement in HRQoL following ART initiation(39). For example, among a random sample of the wider adult population in South Africa, Thomas et al., showed that there was no difference in HRQoL among individuals on ART compared to HIV-negative individuals(24). Collectively, these findings demonstrate the association between ART and higher HRQoL among PLHIV, and the importance of improving ART retention.

We did not observe an independent association between CD4 count or viral load with HRQoL in our study. Although HRQoL scores (utility measures) for PLHIV are often stratified by CD4 count (with lower CD4 counts correlated with lower weights), these estimates were obtained before ART became widely available(6). Indeed, in the United States, more recent utility estimates have been found to be more similar across CD4 count strata and to have a narrower range than pre–ART measures(6). Studies have shown that virally suppressed individuals still experience high levels of symptom-related distress, such as fatigue and energy loss, insomnia, sadness and depression, sexual dysfunction and changes in body appearance(40), which likely reduce their HRQoL. These results support that viral suppression is an essential but not sole end point of HIV care, and there is a need to improve HIV outcomes beyond viral suppression(3, 4).

Our study has several limitations. Our sample did not include FSW living with HIV who were either undiagnosed or diagnosed recently. However, a previous study in South Africa has shown similar HRQoL score between those undiagnosed and those on ART(24). As such, one might be able to approximate the HRQoL score for FSW living with undiagnosed HIV using our estimates of FSW on ART. Our results could be subject to selection bias to the extent that women reached by the sex worker programs and who consented to enroll into the study were different from their counterparts. Nevertheless, it is essential to study HRQoL in marginalized populations such as FSW in high-burden countries. We relied on individuals’ self-report of ART use which might be affected by recall bias. In addition, we used the EQ-5D health state valuations from Zimbabwe, as valuations for South Africa were not available. Our sensitivity analyses using UK valuations produced similar results. Moreover, Zimbabwe value set was obtained in 2000(21). Individual preferences for health states might have changed over time. However, the same value set was commonly used to calculate HRQoL scores across populations in South Africa(24), facilitating the comparison of our estimates against others. Our analyses were based on cross-sectional data and thus cannot be used to infer causality. Caution should be exercised when applying our estimates to FSW populations in different settings.

## CONCLUSIONS

In summary, we evaluated the overall and characteristics associated with HRQoL for FSW living with HIV in South Africa and presented both overall and subgroup-specific estimates of HRQoL score, representing health utility. The findings here reinforce the importance for HIV and sex worker programs to develop and adopt a comprehensive, and human-centered approach, with tailored services designed to address social and structural vulnerabilities including drug use, violence, and stigma to improve HRQoL among FSW living with HIV. In addition, monitoring of HRQoL could provide an actionable measure during program evaluation, and thus inform service adaptation or scale-up to improve the overall well-being among FSW living with HIV.

## Data Availability

The data that support the findings of this study are available from the author, SB, upon reasonable request and approval.

## Declarations

### Ethics approval and consent to participate

The study was approved by the University of the Western Cape Biomedical Research Ethics Committee in South Africa; the Johns Hopkins School of Public Health Institutional Review Board in the United States, and the eThekwini Municipality and KwaZulu-Natal Provincial Departments of Health. Participants provided informed consent prior to enrollment.

### Consent for publication

Not applicable.

### Availability of data and materials

The data that support the findings of this study are available from the author, [SB], upon reasonable request and approval.”

### Competing interests

There are no conflicts of interest to disclose.

### Funding

This study was funded by the National Institutes of Health (R01NR016650).

### Author’s contribution

All listed authors contributed significantly to the work. LW and SM conceptualized and designed the study. CAC, KY, MM, NM, and HM contributed to the data collection. LW and CAC cleaned the data. CC provided critical inputs into data cleaning. LW conducted the data analyses, interpreted the results, and drafted the manuscript. DWD and SM provided critical inputs into the analyses. DWD, CAC, KY, MM, NM, HM, CC, HH, SRS, SB, and SS contributed to the interpretation of results and provided critical manuscript revision. KY, MM, NM, HM, and HH provided important inputs from a local programming perspectives.

#### List of Abbreviations

HRQoL: health-related quality of life
PLHIV: people living with HIV
FSW: female sex workers (FSW)
ART: antiretroviral treatment
QALY: quality adjusted life years
CI: confidence interval
VAS: visual analogue scale
EQ-5D-3L: EuroQoL five dimension, three level

## Additional files

Title of the additional file 1: Health-related quality of life among female sex workers living with HIV in South Africa Appendix.

Format: word doc.

Contents: This additional file includes the following sections and have been cited accordingly in the main text: A1. Details on stigma measures; A2. Details on model fitting and comparison; A3. Supplementary results.

## Notes

### Competing Interest Statement

The authors have declared no competing interest.

### Author Declarations

The study was approved by the University of the Western Cape Biomedical Research Ethics Committee in South Africa; the Johns Hopkins School of Public Health Institutional Review Board in the United States, and the eThekwini Municipality and KwaZulu-Natal Provincial Departments of Health.

