## Appendices for "Health-related quality of life of female sex workers living with HIV in South Africa: a cross-sectional study"

**Appendix**

**Authors**: Linwei Wang^1^, David W. Dowdy^2^, Carly A. Comins^2^, Katherine Young^3^, Mfezi Mcingana^3^, Ntambue Mulumba^3^, Hlengiwe Mhlophe^3^, Claire Chen^2^, Harry Hausler^3^, Sheree R. Schwartz^2^, Stefan Baral^2^, Sharmistha Mishra^1,4,5,6^, on behalf of the Siyaphambili Study team

**A0. Additional details on the study sample**

The current analysis uses baseline data from an adaptive randomized intervention trial (Siyaphambili study) involving FSW in Durban, South Africa. The trial aimed to evaluate the effectiveness of individualized case management and decentralized treatment program to improve viral suppression among female sex workers living with HIV. Cisgender women who were 18 years and older, selling sex as their main source of income, residing in Durban, and diagnosed with HIV at least 6 months prior to study enrollment were eligible.

Study recruitment was conducted by TB HIV Care, a South African nongovernmental organization that has been providing HIV prevention and care for FSW in Durban since 2012. FSW were recruited by peer case managers in the community via mobile van outreach, and from service recipients accessing TB HIV Care services at a drop-in center.

The rationale for inclusion of solely cisgender women: the needs of transgender FSW women living with HIV are important and their experiences, including barriers/facilitators to ART access, use, and retention – especially in the context of case management – may be unique and require specification. And therefore, the focus on individualized case management was to address the barriers and challenges faced by cisgender FSW living with HIV. As such, additional study to measure HRQoL across key populations, including transgender women, will be an important next step.

The rationale for inclusion of individuals >=18 years: sex work under the age of 18 is internationally considered child prostitution and individuals would require parent assent.

The rationale for inclusion of individuals diagnosed with HIV at least 6 months prior to study enrollment: to maximize the generalizability of the study, the study will focus broadly on FSW living with HIV. However, because we wanted the interventions, which are intensive, to focus on women who are most in need of additional support, only those women with at least six months exposure to South African SoC will be eligible for the study and only those not virally suppressed will be randomized to the two intervention arms. By ensuring that a woman has been aware of her HIV status for at least six months, we ensured that those women enrolled into the study had an opportunity to seek out and take up ART treatment and thus if they are not on ART or are not virally suppressed, the SoC package has not been observed to meet their needs. Eligibility will be independent of current or past ART. Individuals on ART, but who were initiated within the past two months will not be eligible because if they are not suppressed it may be an adherence issue or it may be that they just have not had sufficient time on ART to become virally suppressed. Therefore, it is important that potential participants currently on ART have at least two months experience on ART. However, after being on ART for two months or more, the individual may be rescreened for study eligibility should they still be interested in study participation and should study enrollment still be ongoing.

**A1. Details on stigma measures**

We used two sets of stigma measurement tools included in the Siyaphambili study baseline questionnaire to generate the stigma measures in our analyses. The first tool included a total of 27 questions asking about experience with stigma as a female sex worker (FSW) or related to HIV (**Box 1**). The second tool included 3 questions capturing individual’s internalized stigma as a FSW (**Box 2**). Both tools have been used to measure stigma for FSW in other studies^2,3^. Although the tools have been used to measure stigma, there is no consensus on how to summarize the measures across a list of questions. Therefore, we detailed the approach we took below including rationales.

We first categorized each of the 30 questions as a binary variable. For example, if an individual answered ‘yes’ to Q1, it will be coded as 1 (some stigma), and if an individual answered ‘no’ to Q1, it will be coded as 0 (no stigma). For questions related to internalized stigma, e.g., Q28, if an individual answered ‘strongly agree’ or ‘agree’ with the statement, it will be coded as 0 (no stigma), and if an individual answered ‘strongly disagree’, ‘disagree’, or ‘neither agree nor disagree’, it will be coded as 1 (some stigma). If an individual refused to answer or answered ‘don’t know’ or if the answer is missing, it will be coded as missing.

We then summed up the variables within each type of stigma to arrive at three ordinal scale variables: experience with stigma as FSW (range: 0-16), experience with stigma related to HIV (range 0-11), and internalized sex worker stigma (range 0-3). The ordinal scale values reflect the number of questions in which an individual reported stigma; however the interval between the values may not have a continuous scale meaning as some questions are highly correlated (e.g., Q4 and Q5).

We examined the correlations among three stigma variables. The correlation between experience with stigma as FSW and experience with stigma related to HIV was high (Pearson correlation = 0.44); while the correlation between internalized sex workers stigma and the other two stigma variables were low (Pearson correlations were 0.05 and 0.05 respectively). Therefore, we further summed up the two variables regarding experience with stigma to arrive at one variable on experience with stigma related to sex work or HIV (range 0-27) given the high correlation and the fact that our research question did not aim to examine/distinguish the source of stigma.

Finally, we categorized each of the two ordinal variables into 4 levels of stigma: no, slight, moderate, and extreme. For experience with stigma related to sex work or HIV, those who demonstrated no stigma in any of the 27 questions will be defined as having no stigma experience, those who demonstrated some stigma in 1-2 questions will be defined as having slight stigma experience, those who demonstrated some stigma in 3-4 questions will be defined as having moderate stigma experience, and those demonstrated some stigma in 5 or more questions will be defined as having extreme stigma experience. The categorization aimed to capture potential dosing effect associated with the level of stigma, and to balance the size of the categories in the categorical variable. Similarly, for internalized sex workers stigma, those who demonstrated no stigma in any of the 3 questions will be defined as having no internalized stigma, those who demonstrated some stigma in 1 question will be defined as having slight internalized stigma, those who demonstrated some stigma in 2 questions will be defined as having moderate internalized stigma, and those demonstrated some stigma in all 3 questions will be defined as having extreme internalized stigma.

| **Box 1. Experience with Stigma**  "Now I am going to ask you some questions about stigma you have experienced. Please do not feel bad about answering as we will not tell anyone about what you will tell us. Your answers are completely voluntary, and you are free to skip questions if you do not feel comfortable answering them.”  Response: No, Yes, Refuse, Don't know | |
| --- | --- |
| Experiences with stigma (as FSW) | |
| No. | Question |
| 1 | Have you ever felt excluded from family activities because you are a sex worker? |
| 2 | Have you ever felt that a family member made a negative remark or gossiped about you because you are a sex worker? |
| 3 | Have you ever felt rejected by your friends because you are a sex worker? |
| 4 | Have you ever been afraid to seek healthcare because someone might learn that you are a sex worker? |
| 5 | Have you ever avoided seeking healthcare because you were afraid someone might learn that you are a sex worker? |
| 6 | Have you ever felt that you were not treated well in a healthcare center because you are a sex worker? For example: waiting longer, being isolated or misguided, not receiving the quality of care that you deserve, etc. |
| 7 | Have you ever been denied health services or have someone keep you from receiving health services because you are a sex worker? |
| 8 | Have you ever felt that a healthcare worker made negative remarks or gossiped about you because you are a sex worker? Healthcare worker: Doctor, nurse, social worker, etc. |
| 9 | Have you ever felt that a uniformed officer refused to protect you because you are a sex worker? Uniformed officer: Police, soldier, etc. |
| 10 | Have you ever avoided carrying condoms because you thought they might cause you problems from a uniformed officer? |
| 11 | Have you ever seen a uniformed officer take or destroy condoms carried by you or another sex worker? |
| 12 | Have you ever felt a uniform officer harassed or intimidated you because you are a sex worker? |
| 13 | Have you ever been arrested on charges related to prostitution? |
| 14 | Have you ever been afraid to be in public places because you are FSW? |
| 15 | Have you ever felt that you were verbally harassed because you’re FSW? |
| 16 | Have you ever felt that you were blackmailed because you are FSW? |
| Experiences with stigma (HIV) | |
| 17 | Have you ever felt excluded from family activities because you are living with HIV? |
| 18 | Have you ever felt that a family member made a negative remark or gossiped about you because you are living with HIV? |
| 19 | Have you ever felt rejected by your friends because you are living with HIV? |
| 20 | Have you ever been afraid to seek healthcare because someone might learn that you are living with HIV? |
| 21 | Have you ever avoided seeking healthcare because you were afraid someone might learn that you are living with HIV? |
| 22 | Have you ever felt that you were not treated well in a healthcare center because you are living with HIV? For example: waiting longer, being isolated or misguided, not receiving the quality of care that you deserve, etc. |
| 23 | Have you ever been denied health services or have someone keep you from receiving health services because you are living with HIV? |
| 24 | Have you ever felt that a healthcare worker made negative remarks or gossiped about you because you are living with HIV? Healthcare worker: Doctor, nurse, social worker, etc. |
| 25 | Have you ever been afraid to be in public places because you are living with HIV? |
| 26 | Have you ever felt that you were verbally harassed because you are living with HIV? |
| 27 | Have you ever felt that you were blackmailed because you are living with HIV? |

| **Box 2. Internalized stigma as a FSW**  Responses: Strongly disagree, Disagree, Neither agree nor disagree, Agree, Strongly agree, Refusal, Don't know | |
| --- | --- |
| No. | Question |
| 28 | Selling sex is a satisfactory and acceptable way of life for me. |
| 29 | For the most part, I do not care if people know I sell sex. |
| 30 | Selling sex does not make me a lesser person. |

**A2. Details on model fitting and comparison**

*Beta regression models*

As the dependent variable has to be bounded (0,1) in the beta regression model, values of 0 and 1 of health related quality of life (HRQoL) score were transformed by the formula y*=[y(N-1)+0.5]/N^4,5^, where y* is the transformed HRQoL score, y is the original HRQoL score, and N is the sample size. We first examined bivariate relationship between each covariate of interest and the HRQoL score, and grouped some categories of the categorical covariates to minimize Bayesian information criterion (BIC) of the model in the event when additional categories did not provide more information (e.g., we regrouped the 3-level viral load variable into binary (virally suppressed (<50 copies/ml) vs. ≥50). We also compared different link functions and selected the models with the complementary log-log link over logit, log-log and probit links based on minimized BIC. We further compared models with fixed dispersion vs. variable dispersion (covariate effect on both mean and precision parameters) and selected model with fixed dispersion based on the likelihood ratio test comparing nested models.

We included all covariates of interest (with the updated categorization) except two in our multivariable beta regression model as they were defined *a priori*. The two excluded covariates were self-identifying as black given that 97.4% of the study population self-identified as black; and injection drug use, as 4% of the study population reported inject drug use, and this variable is highly correlated with drug use. Finally, we examined several models with interactions, including interactions between antiretroviral treatment (ART) experience (on vs. off ART) and other covariates, separately (one interaction term at a time), and interactions between viral load and CD4 count. We compared the models with interactions against the multivariable model without interaction using likelihood ratio tests to assess if treatment status modified the relationship between other covariates and HRQoL score, and if there was a synergistic effect of both clinical factors on the HRQoL score. Models with any of the interaction terms did not improve the model fitting.

*Alternative regression models*

In order to assess if model specification influenced our results, we alternatively fitted a multivariable linear regression with robust standard error, and a two-part model (given excess extreme values in HRQoL score (HRQoL score equal to 1) ) to identify the determinants of HRQoL score. Both methods have been used in the literature to model EQ-5D HRQoL score, in addition to the beta regression [**refs**]. We found model specification did not modify the list of identified determinants for the primary outcome (HRQoL score) (**Table A2.1; Table A2.2**). We chose beta regression over linear regression based on minimized BIC; and over the two-part model for a more straight-forward interpretation than summarizing two parts.

**Table A2.1.** Multivariable linear regression analyses of factors associated with the EQ-5D health-related quality of life (HRQoL) score of female sex workers living with HIV in Durban, South Africa, 2018-2020.

|  | Average change  in EQ-5D HRQoL score* | | | |
| --- | --- | --- | --- | --- |
| Characteristics | Point | Lower | Upper | P-value |
| Age in years (per decade increase) | **-0.038** | **-0.052** | **-0.023** | **<0.001** |
| Completed secondary school or higher | 0.008 | -0.017 | 0.034 | 0.51 |
| Current on ART | **0.037** | **0.013** | **0.061** | **0.002** |
| CD4<200, cells/mm^3^ | -0.008 | -0.041 | 0.026 | 0.66 |
| Viral suppression (<50 copies/ml) | -0.004 | -0.027 | 0.018 | 0.70 |
| Drug use, past 30 days | **-0.040** | **-0.061** | **-0.018** | **<0.001** |
| Homelessness, past 6 months | -0.015 | -0.039 | 0.008 | 0.20 |
| Experience of physical or sexual violence, ever | **-0.052** | **-0.073** | **-0.031** | **<0.001** |
| Experience with stigma (slight vs no)** | 0.004 | -0.023 | 0.030 | 0.78 |
| Experience with stigma (moderate vs no)** | 0.017 | -0.012 | 0.045 | 0.25 |
| Experience with stigma (extreme vs no)** | 0.003 | -0.028 | 0.034 | 0.84 |
| Internalized stigma (slight vs no)*** | -0.013 | -0.039 | 0.014 | 0.36 |
| Internalized stigma (moderate vs no) *** | **-0.042** | **-0.069** | **-0.014** | **0.003** |
| Internalized stigma (extreme vs no) *** | 0.006 | -0.023 | 0.035 | 0.68 |

*Estimated using linear regression with robust standard error (HC3 type covariance structure); the HRQoL score was calculated using the Zimbabwe general population value set; **HIV or sex work related stigma, ever; details on stigma measures in **Appendix A1**; ***sex work related; details on stigma measures in **Appendix A1**;

Abbreviations: EQ-5D, EuroQoL five dimension; ART, antiretroviral treatment.

**Table A2.2.** Multivariable regression analyses using two part model* of factors associated with the EQ-5D health-related quality of life (HRQoL) score** of female sex workers living with HIV in Durban, South Africa, 2018-2020.

|  | **Part 1: Multivariable logistic regression**  Adjusted odds ratio associated with  perfect health (having HRQoL score of 1; yes vs. no) | | | | **Part 2: Multivariable beta regression among those having HRQoL score <1**  Average marginal change  in EQ-5D HRQoL score | | | |
| --- | --- | --- | --- | --- | --- | --- | --- | --- |
| Characteristics | Point | Lower | Upper | P-value | Point | Lower | Upper | P-value |
| Age in years (per decade increase) | **0.72** | **0.62** | **0.84** | **<0.001** | **-0.024** | **-0.038** | **-0.009** | **0.002** |
| Completed secondary school or higher | 1.17 | 0.87 | 1.56 | 0.30 | 0.000 | -0.029 | 0.029 | 0.99 |
| Current on ART | **1.28** | **0.99** | **1.66** | **0.058** | **0.033** | **0.008** | **0.059** | **0.010** |
| CD4<200, cells/mm^3^ | 0.94 | 0.66 | 1.34 | 0.74 | -0.004 | -0.040 | 0.032 | 0.82 |
| Viral suppression (<50 copies/ml) | 0.81 | 0.62 | 1.06 | 0.13 | 0.017 | -0.009 | 0.044 | 0.20 |
| Drug use, past 30 days | **0.62** | **0.48** | **0.8** | **<0.001** | -0.014 | -0.040 | 0.011 | 0.26 |
| Homelessness, past 6 months | 0.85 | 0.66 | 1.1 | 0.22 | -0.007 | -0.032 | 0.018 | 0.57 |
| Experience of physical or sexual violence, ever | **0.61** | **0.48** | **0.78** | **<0.001** | **-0.031** | **-0.056** | **-0.007** | **0.013** |
| Experience with stigma (slight vs no)*** | 1.12 | 0.83 | 1.51 | 0.47 | -0.002 | -0.032 | 0.029 | 0.91 |
| Experience with stigma (moderate vs no)*** | 1.26 | 0.91 | 1.73 | 0.16 | -0.001 | -0.033 | 0.032 | 0.97 |
| Experience with stigma (extreme vs no)*** | 0.94 | 0.67 | 1.32 | 0.72 | 0.016 | -0.017 | 0.049 | 0.34 |
| Internalized stigma (slight vs no)**** | 0.85 | 0.62 | 1.15 | 0.29 | 0.005 | -0.027 | 0.036 | 0.77 |
| Internalized stigma (moderate vs no) **** | **0.65** | **0.48** | **0.88** | **0.005** | -0.013 | -0.042 | 0.017 | 0.40 |
| Internalized stigma (extreme vs no) **** | 0.98 | 0.7 | 1.37 | 0.91 | 0.019 | -0.015 | 0.052 | 0.28 |

*Chosen as there were excess 1 in the outcome; **the HRQoL score was calculated using the Zimbabwe general population value set; ***HIV or sex work related stigma, ever; details on stigma measures in **Appendix A1**; ****sex work related; details on stigma measures in **Appendix A1**;

Abbreviations: EQ-5D, EuroQoL five dimension; ART, antiretroviral treatment.

**A3. Supplementary results**

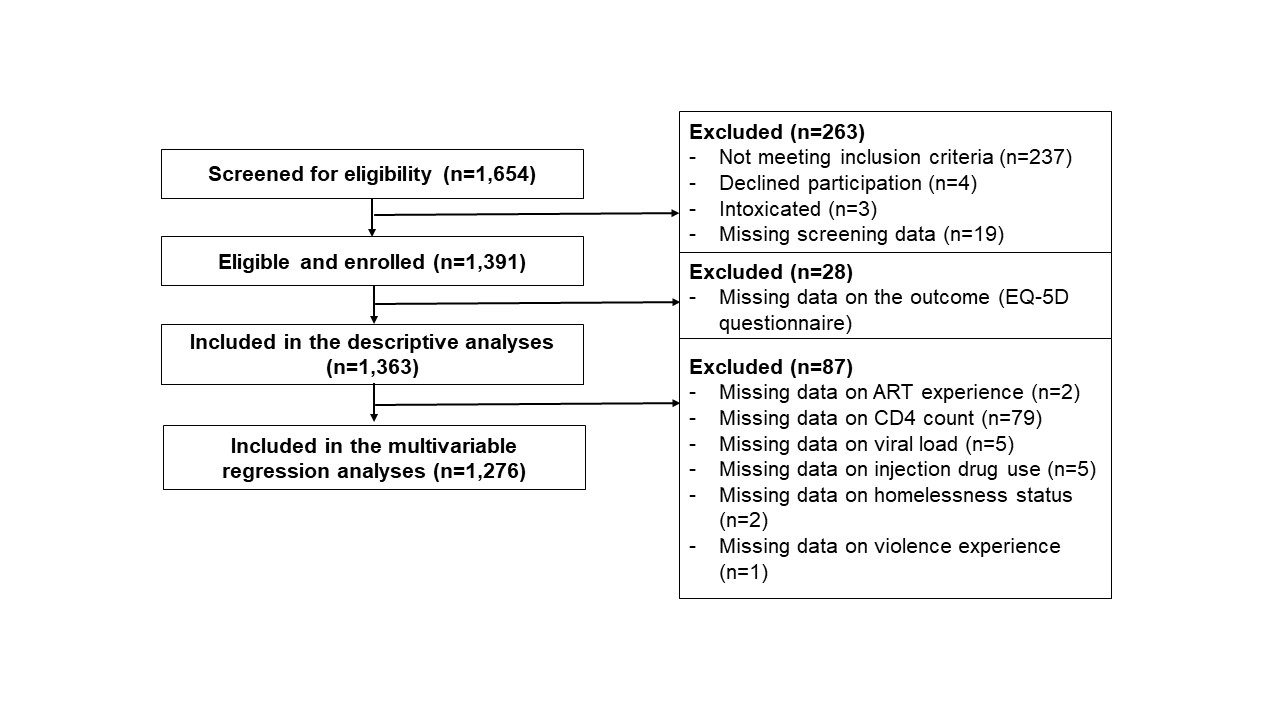

**Figure A3.1.** CONSORT diagram of study sample screened, enrolled, and included in the analyses. Abbreviations: ART, antiretroviral treatment.

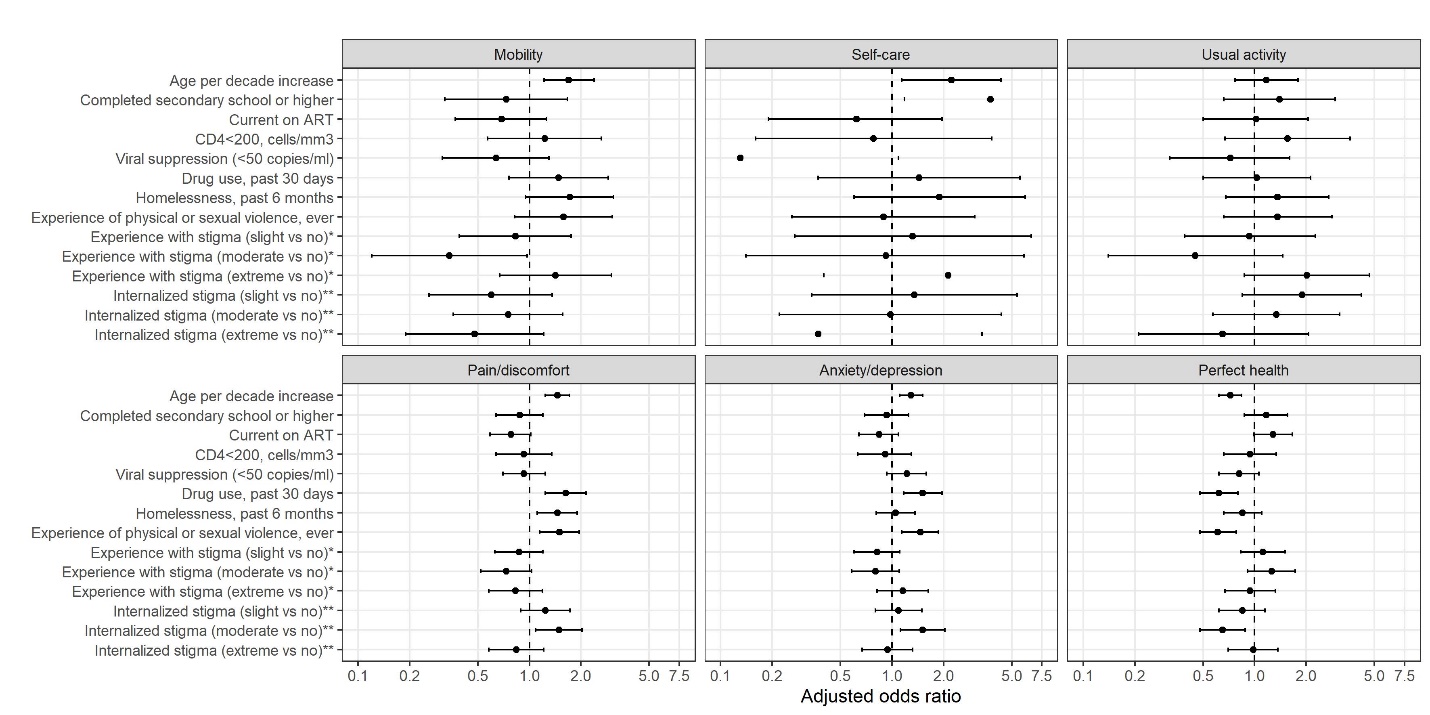

**Figure A3.2.** Multivariable analyses of factors associated with reporting any problem on each EQ-5D-3L domain and perfect health state of female sex workers living with HIV in Durban, South Africa, 2018-2020. Perfect health state is defined as reporting no problem on all five EQ-5D-3L domains. All individuals reporting no problem in self-care and usual activity domains identified as non-black, thus the variable black race was not included in the models for self-care and usual activity. The full range of confidence intervals are not shown in the figure for several variables due to very wide confidence intervals (education, viral suppression, stigma covariates in the model with self-care outcome). *HIV or sex work related stigma, ever; details on stigma measures in **Appendix A1**; **sex work related; details on stigma measures in **Appendix A1**; Abbreviations: EQ-5D, EuroQoL five dimension; ART, antiretroviral treatment.

**Table A3.1.** Comparison of characteristics of female sex workers living with HIV included vs. excluded (due to either missing in outcome or covariates) in the multivariable regression analyses.

|  | **Included** | **Excluded** | **P-value*** |
| --- | --- | --- | --- |
|  | **(N=1276 (91.7%))** | **(N=115 (8.3%))** |  |
| Age in years category |  |  | 0.029 |
| 18-24 | 205 (16.1%) | 27 (26.2%) |  |
| 25-34 | 660 (51.7%) | 53 (51.5%) |  |
| 35-44 | 300 (23.5%) | 19 (18.4%) |  |
| 45-65 | 111 (8.70%) | 4 (3.88%) |  |
| Self-identified as black |  |  | 0.20 |
| Yes | 1243 (97.4%) | 98 (95.1%) |  |
| No | 33 (2.59%) | 5 (4.85%) |  |
| Completed secondary school or higher | |  | 0.79 |
| Yes | 245 (19.2%) | 18 (17.5%) |  |
| No | 1031 (80.8%) | 85 (82.5%) |  |
| ART experience |  |  | 0.067 |
| ART_naive | 166 (13.0%) | 13 (12.5%) |  |
| Currently off ART | 278 (21.8%) | 33 (31.7%) |  |
| Currently on ART | 832 (65.2%) | 58 (55.8%) |  |
| CD4 count, cells/mm^3^ |  |  |  |
| >=500 | 655 (51.3%) | 14 (50.0%) | 0.97 |
| 350-499 | 254 (19.9%) | 6 (21.4%) |  |
| 200-349 | 206 (16.1%) | 5 (17.9%) |  |
| <200 | 161 (12.6%) | 3 (10.7%) |  |
| Viral load, copies/ml |  |  | 0.55 |
| <50 | 478 (37.5%) | 44 (42.3%) |  |
| 50-1000 | 192 (15.0%) | 16 (15.4%) |  |
| >1000 | 606 (47.5%) | 44 (42.3%) |  |
| Drug use, past 30 days |  |  | 0.62 |
| Yes | 789 (61.8%) | 74 (64.3%) |  |
| No | 487 (38.2%) | 41 (35.7%) |  |
| Injection drug use, ever |  |  | 0.19 |
| Yes | 50 (3.93%) | 7 (6.93%) |  |
| No | 1221 (96.1%) | 94 (93.1%) |  |
| Homeless, past 6 months |  |  | 0.012 |
| Yes | 364 (28.5%) | 41 (40.6%) |  |
| No | 912 (71.5%) | 60 (59.4%) |  |
| Experience of physical or sexual violence, ever | |  | 0.087 |
| Yes | 776 (60.8%) | 69 (69.7%) |  |
| No | 500 (39.2%) | 30 (30.3%) |  |
| Experience or anticipation of HIV or sex work stigma, ever* | |  | 0.13 |
| No | 375 (29.4%) | 35 (35.4%) |  |
| Slight | 360 (28.2%) | 33 (33.3%) |  |
| Moderate | 294 (23.0%) | 14 (14.1%) |  |
| Extreme | 247 (19.4%) | 17 (17.2%) |  |
| Internalized sex work stigma** | |  | 0.50 |
| No | 507 (39.7%) | 43 (43.4%) |  |
| Slight | 262 (20.5%) | 24 (24.2%) |  |
| Moderate | 294 (23.0%) | 20 (20.2%) |  |
| Extreme | 213 (16.7%) | 12 (12.1%) |  |

*Fishers exact tests were used; **HIV or sex work related stigma, ever; details on stigma measures in Appendix A1; ***sex work related; details on stigma measures in Appendix A1;

Abbreviations: ART, antiretroviral treatment.

**Table A3.2.** Multivariable analysis of factors associated with EQ-5D health-related quality of life (HRQoL) score (calculated using UK general population value set) of female sex workers living with HIV in Durban, South Africa, 2018-2020.

|  | Average marginal change  in EQ-5D HRQoL score* | | | |
| --- | --- | --- | --- | --- |
| Characteristics | Point | Lower | Upper | P-value |
| Age in years (per decade increase) | **-0.032** | **-0.049** | **-0.016** | **<0.001** |
| Completed secondary school or higher | 0.010 | -0.021 | 0.040 | 0.54 |
| Current on ART | **0.031** | **0.002** | **0.059** | **0.036** |
| CD4<200, cells/mm^3^ | -0.009 | -0.048 | 0.030 | 0.66 |
| Viral suppression (<50 copies/ml) | -0.011 | -0.039 | 0.018 | 0.47 |
| Drug use, past 30 days | **-0.037** | **-0.063** | **-0.011** | **0.006** |
| Homelessness, past 6 months | -0.013 | -0.042 | 0.015 | 0.36 |
| Experience of physical or sexual violence, ever | **-0.044** | **-0.069** | **-0.019** | **<0.001** |
| Experience with stigma (slight vs no)** | 0.007 | -0.025 | 0.039 | 0.67 |
| Experience with stigma (moderate vs no)** | 0.018 | -0.016 | 0.052 | 0.31 |
| Experience with stigma (extreme vs no)** | 0.000 | -0.038 | 0.038 | 1.00 |
| Internalized stigma (slight vs no)*** | -0.013 | -0.046 | 0.020 | 0.44 |
| Internalized stigma (moderate vs no) *** | **-0.039** | **-0.072** | **-0.005** | **0.023** |
| Internalized stigma (extreme vs no) *** | 0.001 | -0.033 | 0.036 | 0.95 |

*Estimated using beta regression with complementary loglog link; **HIV or sex work related stigma, ever; details on stigma measures in **Appendix A1** ***sex work related; details on stigma measures in **Appendix A1**;

Abbreviations: EQ-5D, EuroQoL five dimension; ART, antiretroviral treatment.
